# Persistent Homology and Gabor Features Reveal Inconsistencies Between Widely Used Colorectal Cancer Training and Testing Datasets

**DOI:** 10.1101/2025.04.07.25325392

**Authors:** Daniel Brito-Pacheco, Riad Ibadulla, Ximena Fernández, Panos Giannopoulos, Constantino Carlos Reyes-Aldasoro

## Abstract

Recent work on computer vision and image processing has relied substantially on open datasets, which allow for an objective comparison of techniques and methodologies. In the area of computational pathology and, more specifically, on colorectal cancer, the dataset NCT-CRC-HE-100K, which consists of 100,000 patches of human tissue stained with Haematoxylin and Eosin has been widely used as a training set for deep learning studies. The patches are grouped into 9 classes of tissue (adipose, background, debris, lymphocytes, mucus, smooth muscle, normal colon mucosa, cancer-associated stroma, colorectal adenocarcinoma epithelium). The set is released with a separate set (CRC-VAL-HE-7K) of 7,180 patches that is commonly used for testing. In this work, features were extracted from both sets first with Persistent Homology, then, with Gabor filters to reveal that the training set presents a rather different distribution from the testing set. Namely, the distribution of features in the 7K-set presents a much higher class overlap than those in the 100K-set, which would imply a much higher separability in the testing set than in the training set.

## 1 Introduction

The development and success of deep learning has relied on the existence of large sets of labelled data, in addition to high computational power provided by hardware like graphical processing units. Medical data usually requires labelling by experts, like radiologists, cytologists and pathologists, all of which have experienced shortages in recent years [1,16,18]. Thus, open datasets, such as those provided in *challenges* through websites such as Grand-Challenge (https://grand-challenge.org/), which, in some cases, are related to conferences like ISBI (IEEE D. Brito-Pacheco et al. International Symposium in Biomedical Imaging) or MICCAI (Medical Image Computing and Computer-Assisted Intervention) are welcome by the community. In many cases, the datasets are linked to a competition where algorithms or results are evaluated with certain criteria, e.g. accuracy or Jaccard index, and then ranked and placed in leaderboards. The reproducibility, interpretation and ranking aspects of some challenges have been scrutinised as rankings can vary depending on certain factors, e.g., rank and aggregate v. aggregate and rank, mean v. median, rank with Hausdorff distance (HD) v. rank with HD95, etc. [14]. Another important factor is the test data used for validation.

In this paper, a commonly used dataset (NCT-CRC-HE-100K [11,12]) is scrutinised. The dataset is commonly used to train deep learning architectures for the classification of histological images of colorectal cancer stained with Haematoxylin and Eosin (H&E). The dataset consists of 100,000 patches of human tissue stained with H&E of healthy and cancerous tissues grouped into 9 categories: adipose, background, debris, lymphocytes, mucus, smooth muscle, normal colon mucosa, cancer-associated stroma, colorectal adenocarcinoma epithelium, which are normally used for training, and a separate set (CRC-VAL-HE-7K) of 7,180 patches is commonly used for testing [13, 19, 21]. Concerns have already been raised about these datasets. In [9], the authors identified biases in the classes by performing classification experiments using simple models that focused solely on the colour of the images and obtaining satisfactory results. The authors also managed to identify compression artifacts that show up in different concentrations by class, leading to high classification accuracies.

This paper focuses onfeatures of the patches that are not related to colour, namely texture and structure. By using Persistent Homology and Gabor filters it was observed that these two datasets were not equivalent in the separability of the classes, revealing further issues with the datasets. A visual analysis of the patches was also performed and the effects of normalisation on them was shown. Using these analyses, light is shed on the difference in qualities of the datasets, with CRC-VAL-HE-7K (7K-set) having a higher separability than NCT-CRC-HE-100K (100K-set). Additionally, an experiment was carried out in which a random forest was trained on the 100K-set and tested on the 7K-set before reversing the roles of the sets to train on 7K-set, and tested on 100K-set. This helped to confirm the differences between the sets and show the implications of training and testing on different-quality datasets.

## 2 Background: Persistent Homology

Persistent Homology (PH) is an important tool that belongs to the mathematical field of Topological Data Analysis (TDA), which allows to extract topological features from datasets and create statistical models. This tool has already seen applications in biomedical imaging [6]. Since PH and TDA are not commonly used in Computer Vision and Image Processing areas, a short review will be presented. For a comprehensive introduction to PH, the reader is referred to [8].

### 2.1 Components and Holes

PH analyses an image or a space to find its *components* and *holes*. A (connected) *component* is the set of a space where elements are connected to each other. In the case of an image, a component would correspond to a group of pixels that are adjacent to each other. A *hole* corresponds to a region that is completely surrounded by a single component and does not belong to that component. PH essentially tracks how the number of components and holes change as the conditions on the space or image change. Fig. 1 (a) illustrates six 2D spaces (binary images) with different numbers of components (1,2,3,3,2,1) and holes (0,0,0,1,1,0). These cases can also be understood as a process from left to right where components and holes are born (appear) and die (disappear). This is illustrated in Fig. 1 (b), (c) where the birth of each is indicated with a vertical black line, and the death is indicated with a vertical grey line with a red cross.

**Fig. 1.**
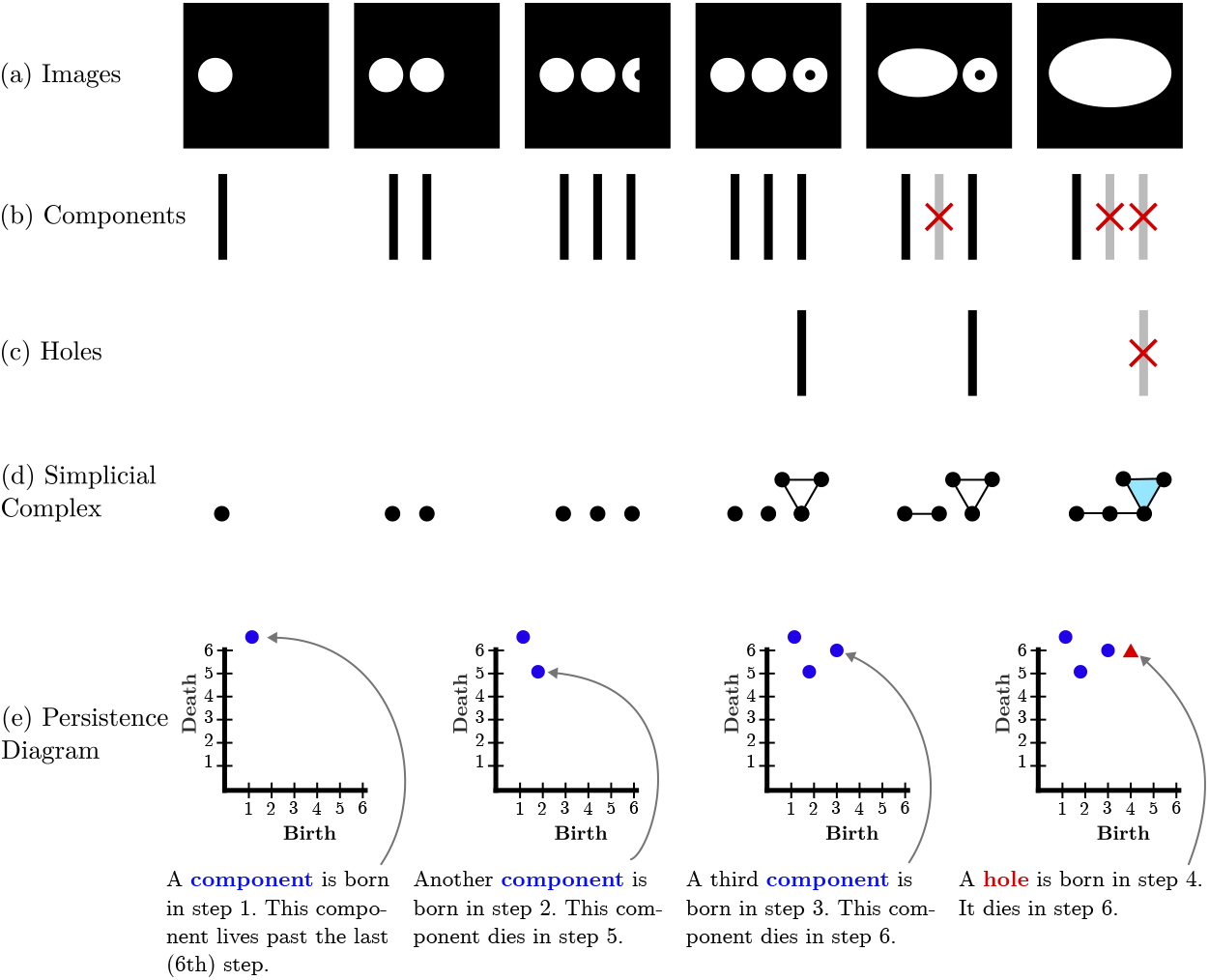
Illustration of the main concepts of Persistence Homology: births, death, holes and components. (a) A sequence of six binary images (b) A black bar corresponds to the birth of a component and, a red cross and a grey bar indicates a death. (c) The birth and death of holes. (d) Representation of the components as a simplicial complex: A point corresponds to a component, except if there is a hole, in which case a cycle of 3 points and 3 edges are added. The cycle is filled with a triangle when the hole dies. (e) Step-by-step formation of the persistence diagram.

### 2.2 Simplicial Complexes

Simplicial complexes are an abstraction from the components and holes previously described. Specifically, an *n-simplex* is an *n*-dimensional generalisation of a triangle. A 0-simplex is a point, a 1-simplex is an edge that connects 2 points, a 2-simplex is a triangle that connects 3 points, a 3-simplex is a tetrahedron, etc. A simplicial complex *S* is a collection of *n*-simplices such that their *n-1* - dimensional faces are also in *S*. This means, for example that if there is an edge *e* in *S*, then the two vertices at the ends of *e* also have to be present in *S*. In the context of PH, simplicial complexes are used as a way to represent a more complex object topologically as illustrated in Fig. 1 (d). The topological information of the holes and components can be represented simply as a collection of vertices, edges and triangles. From left to right: A vertex appears when the first white component appears, a second vertex is added in the second column. a third vertex is added in the third column, a hole appears which is represented by a cycle of edges and their vertices, two components merge into one represented by an edge in row (d) (the component that was born last has died at this point), the hole is filled up; represented by adding a blue triangle in row (d) (the hole has died at this point).

It is important to highlight two points. First, a cycle of edges is different from a triangle. In the first case, the cycle has a hole, whereas a triangle has no hole. Second, a component in a simplicial complex is any union of vertices, edges, triangles, tetrahedron that are touching. A sequence of *n* simplicial complexes *S*_*i*_ is called a *filtration* if *S*_1_ ⊂ *S*_2_ ⊂*S*_3_ ⊂ … ⊂ *S*_*n*_. In Fig. 1 the sequence of simplicial complexes in row (d) makes a filtration. It is also worth mentioning that, in the literature on PH, the formal definition of filtrations applies to sequences of topological spaces, not just simplicial complexes.

### 2.3 Birth, Death, and Persistence Diagrams

*Persistence Diagrams* are a way to encode invariants about a filtration. Particularly, the points when a new topological feature appears, as well as when it disappears in the filtration. Alluded to before, at the step when a component or hole first appears, it is said to be “born”. Analogously, at the step when a component or hole disappears from the filtration, it is said that it “dies”. This information can be encoded in a scatter plot where the horizontal coordinate shows the birth of the component or hole, and the vertical coordinate shows the death of the component or hole as illustrated in Fig. 1 (e).

It is important to note that holes die by being filled in, but components die by merging into each other. When two components merge, there is a choice to be made as to which component dies. Typically, the eldest component of the merger will survive.

### 2.4 Level-Set Filtration

A very common way to get a filtration from a greyscale image is through the level-set filtration, which is illustrated in Fig. 2. First, a greyscale image (Fig. 2 (a)) is thresholded at decreasing intensities to produce a series of binary images (Fig. 2 (d)). Second, a vertex is added at every white pixel, an edge between the vertices if the corresponding pixels neighbour each other, and a triangle if three edges form a cycle (Fig. 2 (e) and Fig. 2 (f)). For completeness, Fig. 2 (b) shows the persistence diagram computed from the filtration of simplicial complexes. There are other methodologies of filtration, which will not be covered in this paper and the reader is referred to [7].

**Fig. 2.**
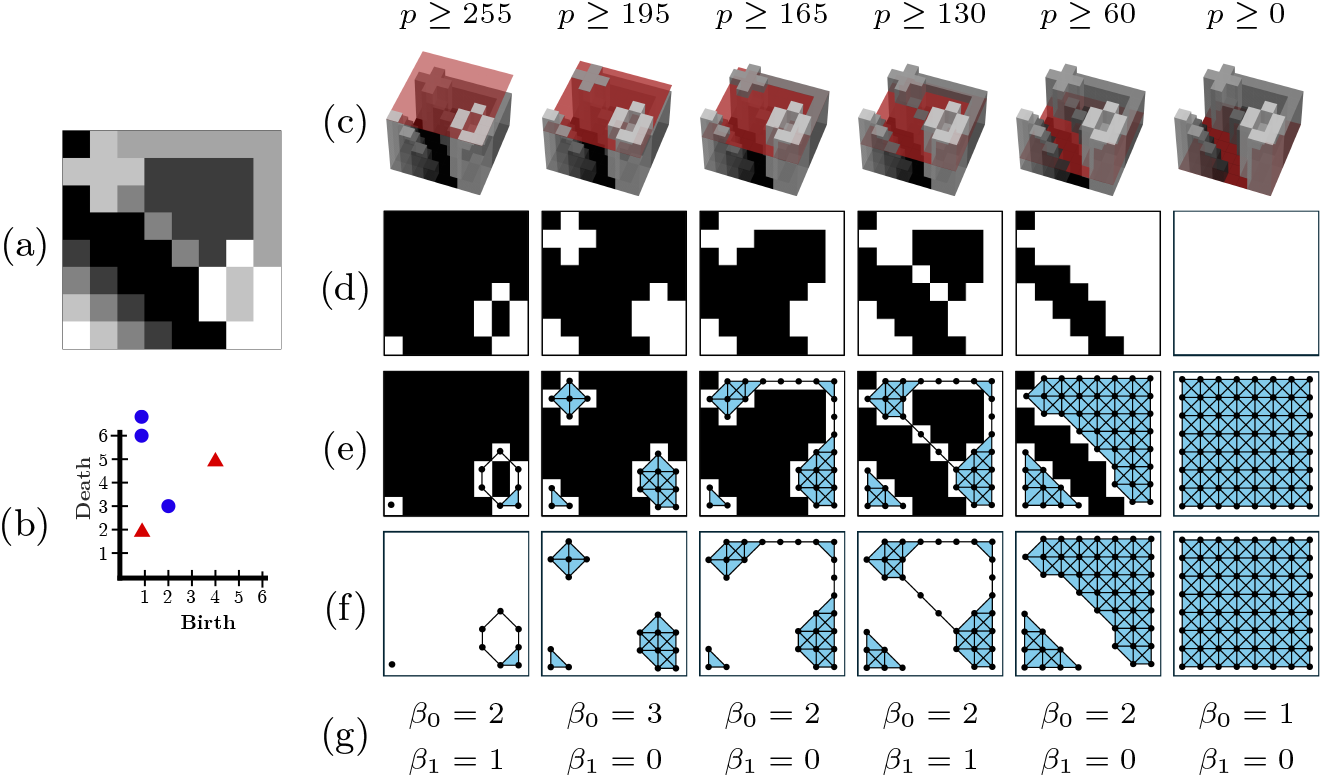
Illustration of the filtration and PH calculation.(a) Original greyscale image in the range [0, 255]. (b) Persistence diagram. (c) 3D representation of the greyscale and a threshold. (d) Binary images of pixels above the threshold. (e) Filtration overlaid on the binary images. A vertex is placed at each white pixel. Edges are added between neighbouring pixels. A triangle is added when cycles of three edges are formed. (f) Filtration with pixels removed. (g) Betti numbers.

## 3 Materials

In this work, two datasets of colorectal cancer slides stained with H&E were used: 100K-set and 7K-set, which contain 100,000 and 7180 images, respectively [11]. Both sets contain images of nine different classes: ADI: adipose tissue; BACK: background; CRC: colorectal cancer; DEB: debris; LYM: lymphocytes; MUC: mucus; MUS: smooth muscle; NORM: normal colon mucosa; STR: cancer-associated stroma; TUM: colorectal adenocarcinoma epithelium (Fig. 3).

**Fig. 3.**
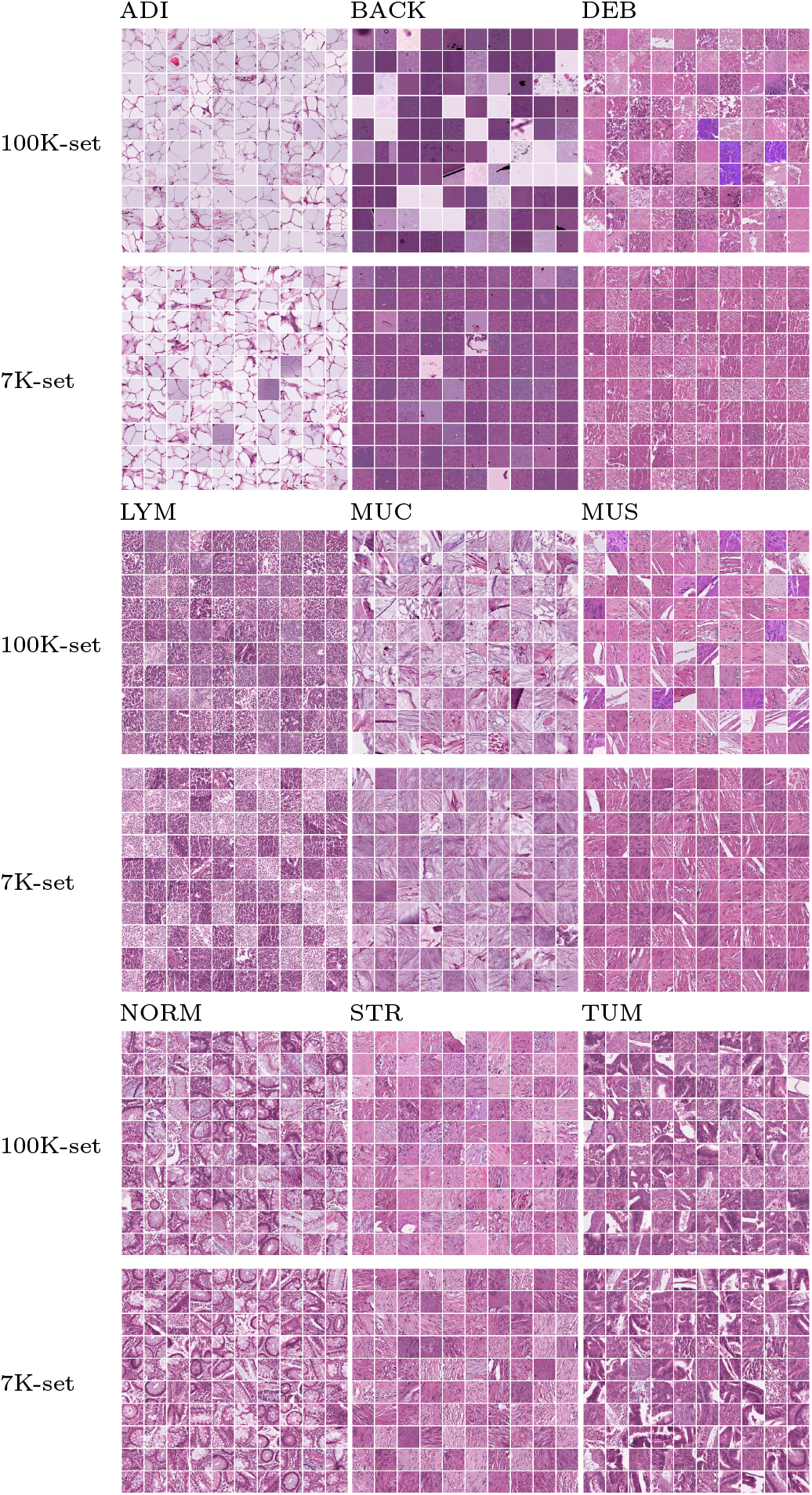
Illustration of the datasets with 100 sample patches per class from each set. By class: NCT-CRC-HE-100K images are shown above, normalized CRC-VAL-HE-7K images are shown below. ADI: adipose tissue; BACK: background; CRC: colorectal cancer; DEB: debris; LYM: lymphocytes; MUC: mucus; MUS: smooth muscle; NORM: normal colon mucosa; STR: cancer-associated stroma; TUM: colorectal adenocarcinoma epithelium.

The 100K-set’s images were explicitly stated to be normalised using Macenko’s method in the paper where the set is introduced [12], but the 7K-set was not explicitly stated to be normalised. Upon closer inspection, it was found that the 7K-set had not been normalised. With the same reference image as the 100K-set, the 7K-set was normalised using Macenko’s method on a per-patch basis before any further analysis was performed (Fig. 4).

**Fig. 4.**
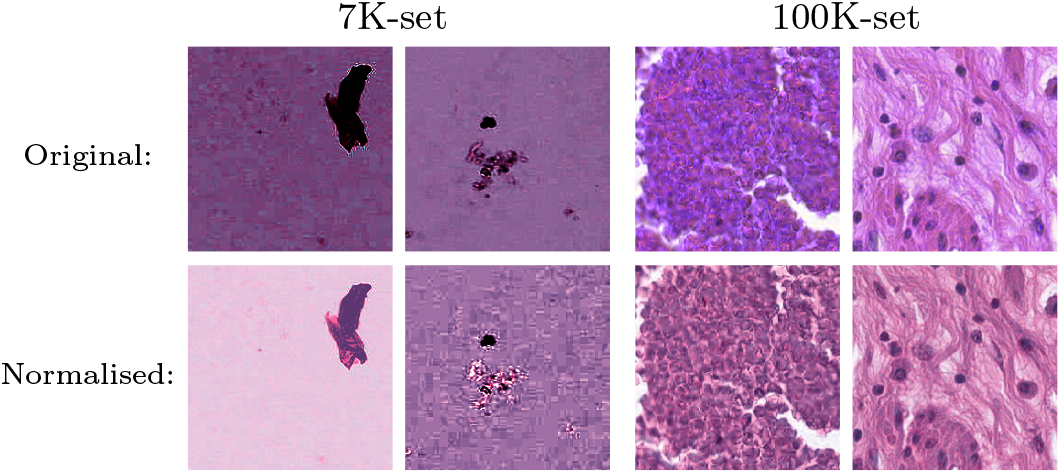
Effects of normalisation. Left: two BACK patches from the 7K-set; one presents a large dark spot causing the rest of the normalised image to become extremely bright, while the other stays relatively uniform. Right: two very purple patches (from DEB and MUS classes), when normalised look visually more like the rest of the set.

## 4 Methods

Two types of features were computed from the images from the train and test datasets. Topological features were obtained from the level-set filtration previously introduced. For comparison purposes, features using Gabor frequency filtering were also calculated. All the features were normalised to a range of 0-100. These methodologies are described below.

### 4.1 Topological Features

Topological features were calculated from images by the following process. First, the colour image was converted to greyscale. Then, a 5 × 5 median filter was applied to the greyscale image to reduce noise and make regions of similar intensities smoother. Next, the persistence diagram was calculated using level-set thresholds in the range [0,255]. Fig. 5 shows some example patches from the 100K-set together with the persistence diagram generated through this process. There is always exactly one component that makes it to the end of the filtration and, in strict mathematical notation, its death is given as ∞. This was also the value returned by the GUDHI [15] package used to compute the diagram. For simplicity, the point corresponding to this component was discarded from the persistence diagram before calculating the following features: number of components/holes, mean birth of components/holes, mean death of components/holes, standard deviation of the births of components/holes, standard deviation of the deaths of components/holes, mean persistence of components/holes, median persistence of components/holes, standard deviation of the persistences of components/holes, minimum birth of components/holes, maximum birth of components/holes, minimum death of components/holes, maximum death of components/holes, range of births of components/holes, range of deaths of components/holes, 1^st^, 5^th^, 25^th^, 50^th^(median), 75^th^, 95^th^, 99^th^percentiles of births of components/holes, 1^st^, 5^th^, 25^th^, 50^th^(median), 75^th^, 95^th^, 99^th^percentiles of deaths of components/holes. The ratio between the number of holes and the number of components was also calculated and added to the list of features, for a total of 57 topological features from each persistence diagram.

**Fig. 5.**
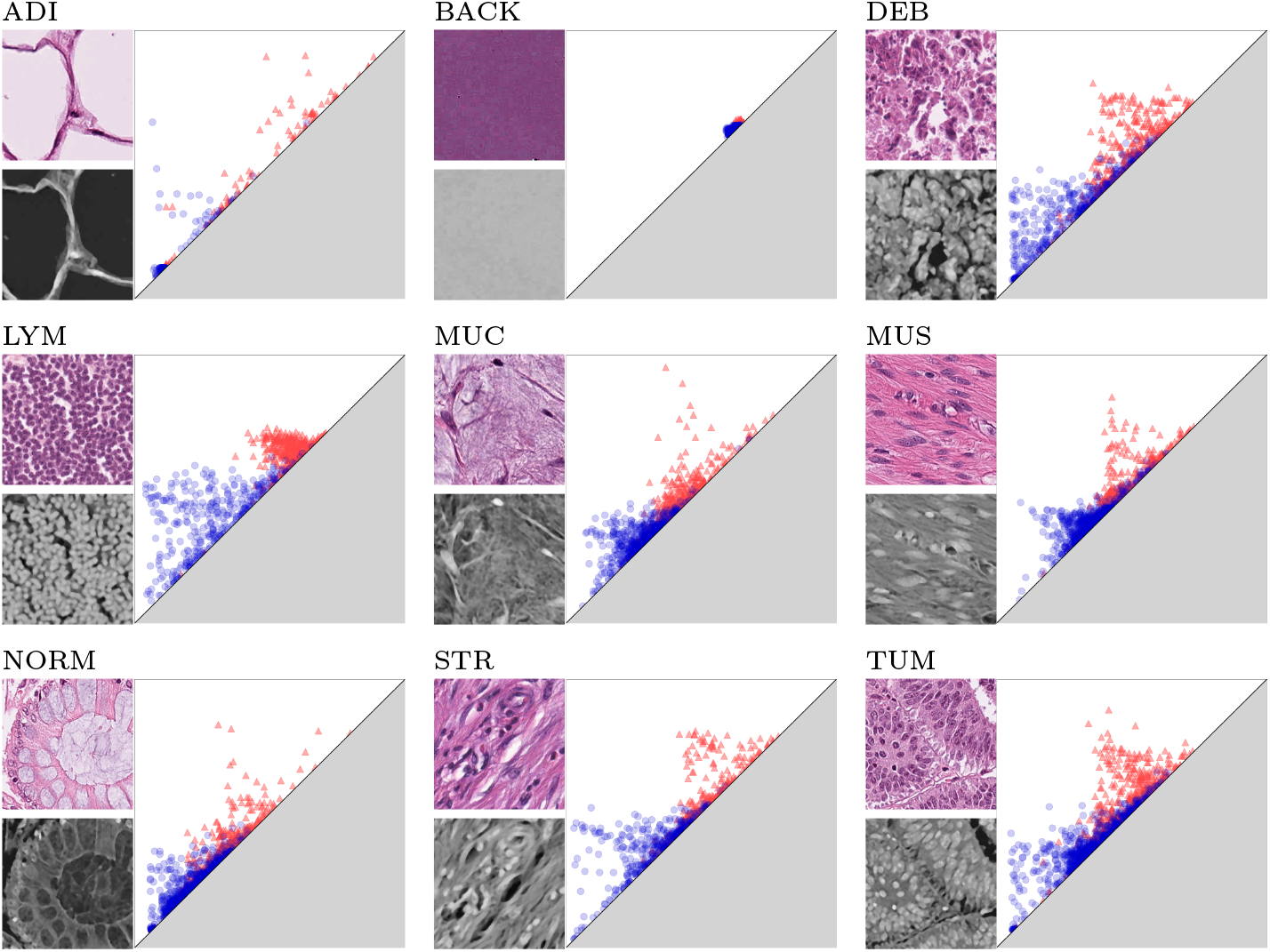
Illustration of the persistence diagrams of the histological tissues from the 100K-set. One representative patch from each of the classes (ADI, BACK, DEB, LYM, MUC, MUS, NORM, STR, TUM) is converted to greyscale and inverted. Noise is removed on the greyscale image by applying a 5 × 5 median filter. A persistence diagram is calculated from the smoothed greyscale image where blue circles are components and red triangles are holes. The distribution of the scatterplots in the persistence diagram capture differences in the textures of different tissues. ADI: adipose tissue; BACK: background; CRC: colorectal cancer; DEB: debris; LYM: lymphocytes; MUC: mucus; MUS: smooth muscle; NORM: normal colon mucosa; STR: cancer-associated stroma; TUM: colorectal adenocarcinoma epithelium.

### 4.2 Gabor Features

Gabor filters are filters which operate on an image through convolution. They can be described as made up of a spatial frequency and orientation within a two-dimensional Gaussian envelope. For an in-depth explanation of how Gabor filters are used, the reader is referred to [17]. Each Gabor filter is defined uniquely by a direction, a frequency, and the standard deviations in the horizontal and vertical coordinates. 36 different Gabor filters of varying directions, frequencies and standard deviations were generated [20]. To compute features using Gabor filters, each image was converted to greyscale and then convolved with the Gabor filters. This yields a filtered greyscale image, from which the mean pixel intensity and variance of pixel intensities were computed. The effects that different directions and frequencies for the Gabor filters have on the filtered image are shown in Fig. 6.

**Fig. 6.**
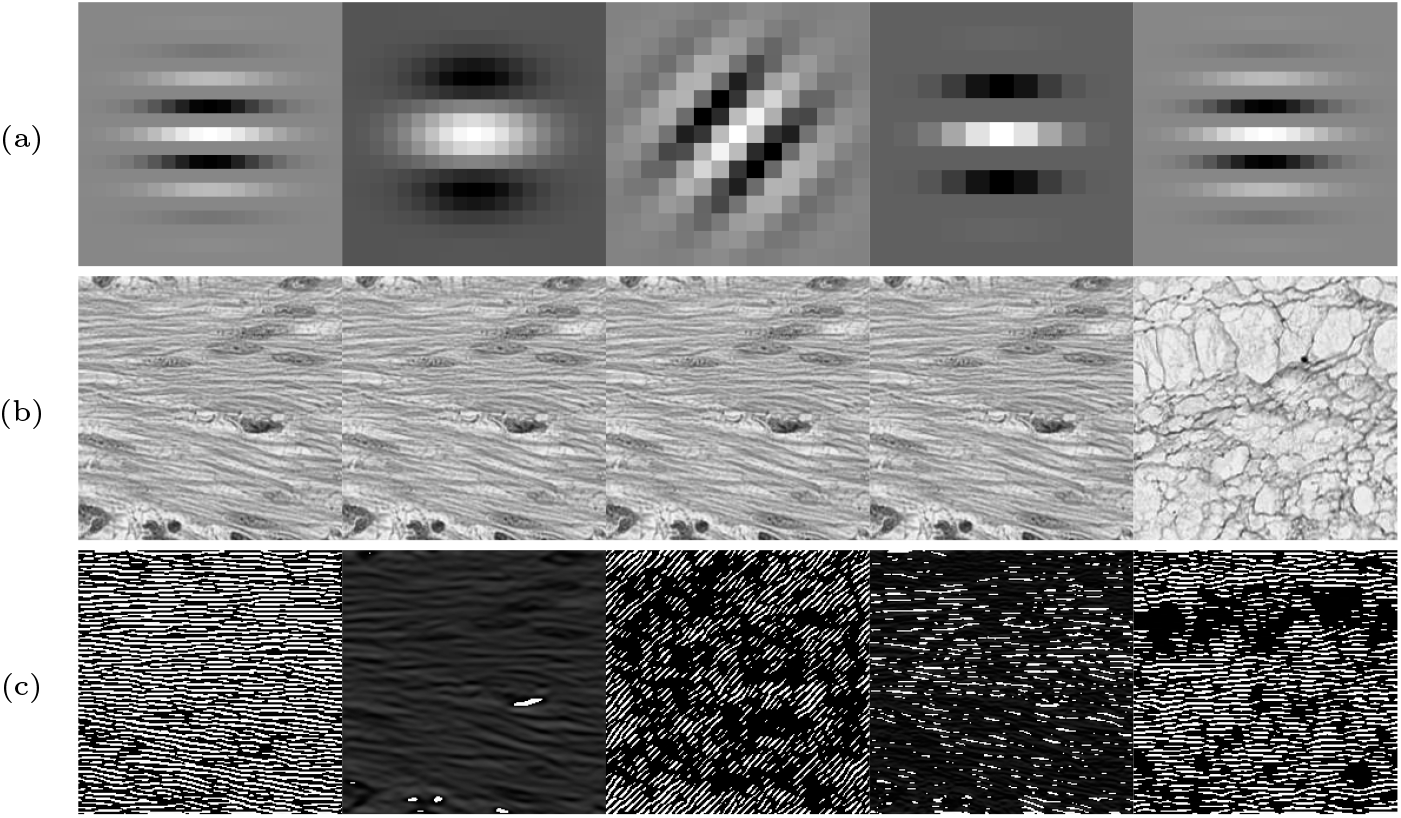
Illustration of Gabor filters. (a) Gabor filters (b) Sample patches converted to greyscales (c) Filtered results.

### 4.3 Random Forest

Random forests are a supervised classification model introduced in 2001 by L. Breiman [4]. It is based on the much older decision tree classifiers [5]. In broad terms, a random forest classifier is created by building multiple decision trees and combining their outputs: the algorithm creates many subsets of the training data by randomly sampling with replacement (this is called “bootstrapping”), then each subset (bootstrapped set) is used to train a single decision tree. Additionally, not all features are considered for each tree - only a random subset of them. For classification, the label assigned to a new sample is given by majority voting from all the decision trees in the forest. A comprehensive overview on random forests can be found in [2].

A random forest model was trained on subsets of different sizes (*n* =100, 250, 500, 750, 1000, 2500, 5000, 7500, 10000, 20000) of the 100K-set and tested on the complete 7K-set, using only topological features, only Gabor features, and the combination of both (Combined features). The model was fit using 100 estimators (decision trees) and a maximum depth of 100 nodes for each estimator. The criterion used to build the decision trees was the Gini index.

The accuracy on the 7K-set was calculated by taking the ratio of correctly classified samples to the total number of samples in the set. The *out-of-bag* (OOB) score of each model was also calculated [3]. As mentioned previously, the model works by bootstrapping the dataset many times; i.e. randomly sampling with replacement many times. Then, decision trees are built using the boot-strapped sets. The OOB score is calculated by classifying the samples which are not included in the bootstrapped sets for each tree. In other words, if a sample is not part of the bootstrapped set used to build a particular tree, the sample is labelled by that tree, and it is checked whether or not the assigned label is correct. The OOB score is a popular way to estimate how well a random forest will generalise, as for large samples it approximates a *k*-fold cross-validation estimation [10]. For both the accuracy and OOB score, a correct classification was considered any sample that was assigned the correct tissue label. When a sample was assigned a different class, it was considered a misclassification.

## 5 Results

The t-SNE visualisations reveal clearly that the separability of the classes is greater in the 7K-set than in the 100K-set for the topological features (Fig. 7(a)), Gabor features (Fig. 7(b)) and Combined features (Fig. 7(c)). To emphasise the separability, 2D Gaussian distributions were fitted to the distributions of the points per class, and the equation of the ellipse containing the area 1.5 standard deviations away from the mean was calculated. Whilst the ellipses for 4 classes overlap substantially in the 100K, MUS and MUC (red and purple) are quite separate from NORM and TUM (green and yellow) in the 7K.

**Fig. 7.**
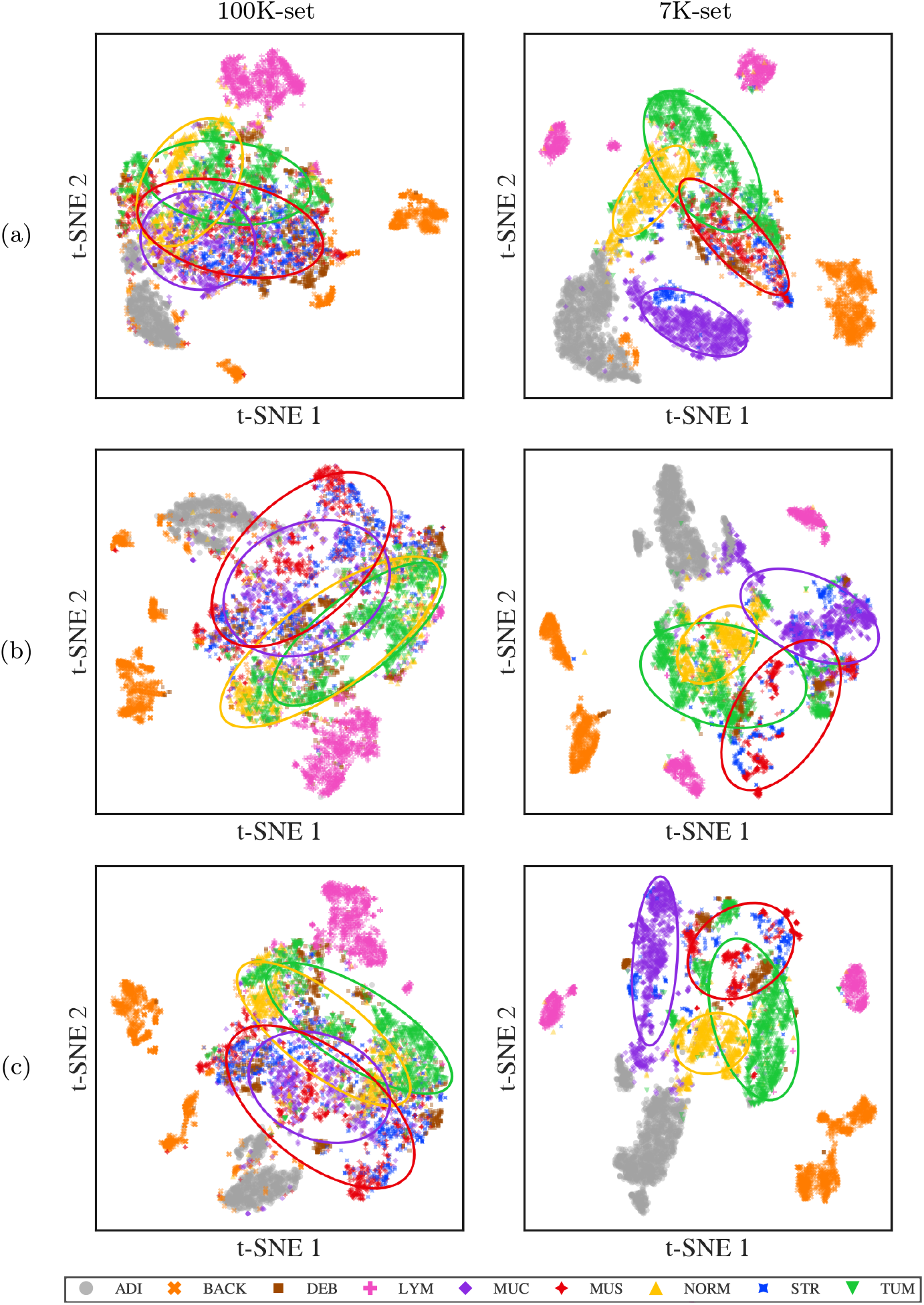
t-SNE visualisation of the samples by feature type and set they belong to. (a) Visualisations created from topological features. (b) Visualisations created from Gabor features. (c) Visualisations created from Combined features.

To confirm the separability of classes between sets, Random Forests were trained with an increasing number of topological, Gabor and Combined features and then tested on the 7K-set (Fig. 8). The results of the OOB score and accuracy follow similar patterns and stabilised between 5,000 and 10,000 samples, which suggested that around 7,000 samples were sufficient to obtain good results.

**Fig. 8.**
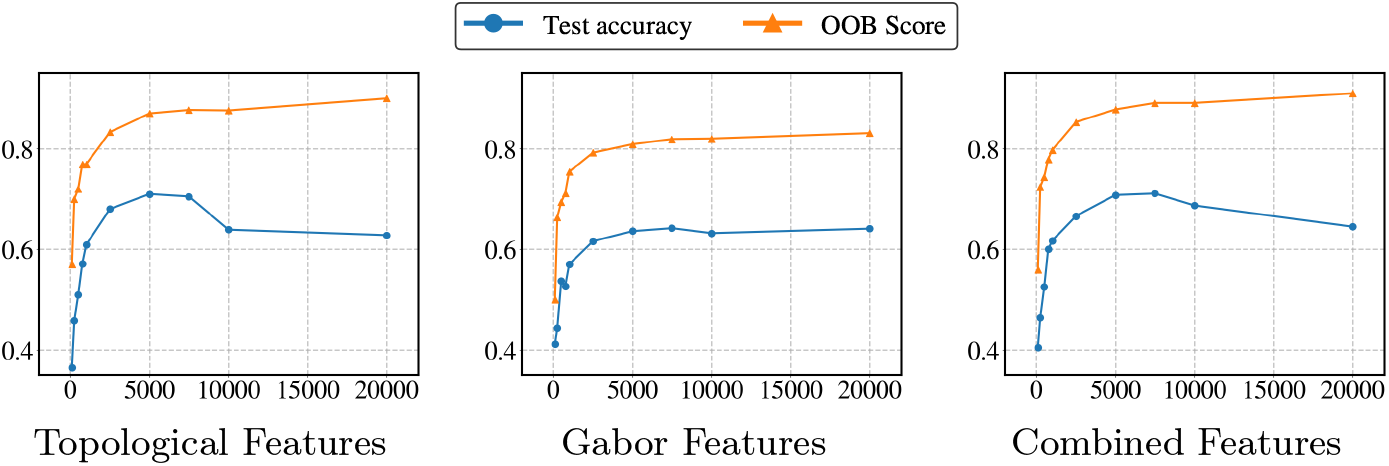
Accuracy on the 7K-set and the OOB score when a Random Forest model is trained on random samples of differing sizes from the 100K-set and using topological, Gabor or combined features.

Next, the number of samples of the 100K-set were restricted to 7,180 to perform a reverse experiment: train on the reduced 100K-set and classify the 7K-set and then train on the 7K-set and classify the reduced 100K-set. The results confirmed the higher separability of the 7K-set, reaching 0.96 OOB-Score but only a 0.55 when the 100K-set was classified with training on the 7K-set (Table 1). On the other hand, when training on the 100K-set, the OOB-Score was lower (0.88) and the classification of the 7K-set was 0.74.

**Table 1.** Effects of training on the different sets. The model was trained on the combined features using 7180 samples from the 100K-set and tested on the 7K-set, then trained on the 7K-set and tested on the 7180 samples from the 100K-set. Note that training on 100K-set and testing on 7K-set yields a much higher Test Accuracy but lower OOB-Score than the reverse case.

| Training set | Test Accuracy | OOB-Score |
| --- | --- | --- |
| 100K-set | 0.6926 | 0.8840 |
| 7K-set | 0.5618 | 0.9642 |

## 6 Discussion

Concerns about the bias in the colour profile of the classes and improper handling of the images 100K-set and the 7K-set had been highlighted [9]. However, up to the best knowledge of the authors, the differences in separability of classes of these important datasets had not been discussed. These were demonstrated with topological and Gabor features, which were used to extract textural and structural properties from the data, that is, properties unrelated to colour.

First, the problems related to **brightness** previously noted in [9] were confirmed. The intensity profiles of the BACK class in the 100K-set go to the extremes; very bright or very dark (Fig. 3). In contrast, the patches in the BACK class in the 7K-set have more uniform intensities. The few patches that are brighter almost always contain a very dark region, these are most likely due to issues of **normalisation**. Macenko’s normalisation seems to have this effect when there is a dark region in the image.

Second, there were problems of **hue**, which are visible in the DEB and MUS classes of the 100K-set. In the DEB class, there are four patches that are visually more purple/violet (hue values around 270-280) than the majority, which are visually closer to pink/magenta (hue values around 300-320). In fact, it was found that the average hue of the four DEB patches was 277, while the average hue of the non-faulty patches was 312. These variations were also present in the MUS class. These differences in colour seem to suggest that the 100K-set has not been properly normalised using Macenko’s method.

Third, issues of **curation and labelling** were detected. The 100K-set shows signs of less consistent curation and labelling when compared to the 7K-set. For example, MUS patches of the 100K-set show white areas (possibly background). This will be further analysed below.

Fourth, there may be differences in **cell populations**, specifically there seem to exist two different types of LYM pathces in the 7K-set. An initial observation of these patches suggests that the difference arises from the brightness of the patches. However, a closer inspection suggests that there is also a difference in the sparseness and size of the lymphocytes (Fig. 3).

Fifth, the **differences in separability of classes** between the 100K-set and the 7K-set became evident with the extraction of topological and Gabor features and the visualisation with t-SNE (Fig. 7), and the previous visual observations were confirmed. Four classes 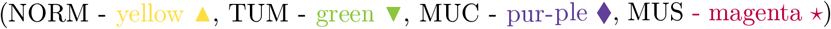 were highlighted with ellipses as previously described. Whilst in the 100K-set, these four classes partially overlap in the topological, Gabor and combined features, in the 7K-set, these appear comparatively separated, especially MUC in the topological and TUM and NORM from MUS and MUC in Gabor and combined. Similarly, the ADI 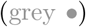 and the BACK 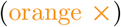 are better separated in the 7K-set. For the topological features, the class BACK is strongly clustered in the 7K-set with just a few elements close to the ADI class, whilst in the 100K-set there are several clusters and the ADI class is surrounded by elements of BACK. For Gabor features, the BACK in 7K-set is totally separated from the ADI class but in the 100K-set again many elements overlap and presents more clusters.

As noted previously, the LYM class in the 100K-set always appears as a single cluster (pink crosses), whilst in the 7K-set always appears as two or even three large and distinctly located clusters (Fig. 7(a-c)). A smaller group is also visible and close to the other classes suggesting that there may be a third type of LYM patches not immediately distinguishable in Fig. 3.

To quantify the effects of the separability of classes, the following experiments were conducted: (1) Train on 7,180 samples from the 100K-set and classify the 7K-set, (2) train on the 7K-set and classify 7,180 samples from the 100K-set (Table 1). When trained on the 7K-set, a random forest model obtained a higher OOB-Score (0.9642) than when trained on 7180 samples of the 100K-set (0.8840). Yet, when the opposite set was classified, the results of the model trained on the 7K-set were far lower (0.5618) than those trained on the 100K-set (0.6926). Both of these results imply a higher separability of classes in the 7K-set.

Separability of classes can come from many factors: intrinsic differences in the classes (i.e. different textures, colours, or shape). However, it is to be expected that two sets of images treated in a similar manner should show the same degrees of separations between their classes. Some prominent factors can potentially influence the observed differences in separability between the sets, mainly differences in preprocessing and handling of the images. In the case of these two data sets, the most probable causes are problems of normalisation and problems of curation and labelling. In the case of normalisation, some patches show evidence of not having been properly normalised when compiling the dataset. This is especially noticeable in Fig. 3 when looking at the DEB and MUS classes and the effects shown in Fig. 4. An example of problems with curation and labelling is that MUS patches have a seemingly more even texture than the patches in the 100K-set: some of these patches seem to show more white areas (possibly background) than actual tissue (Fig. 3). However, other factors such as the probable existence of three different populations of lymphocytes could suggest that there may be intrinsic differences related to the nature of the tissue (healthy/diseased) in one particular patient.

To summarise, greyscale versions of images from two datasets (100K-set and 7K-set) were used to extract features from the corresponding persistence diagrams and the Gabor-filtered versions of the images. The features were used to train random forest models (Fig. 8) and compare the effects of training on one of the sets and testing on the other, then reversing the roles (Table 1). The difference in accuracies and OOB-Score of the experiment, together with a visual analysis of the images (Fig. 3) and a t-SNE visualisation of the classes reveal inconsistencies between the datasets. These inconsistencies go beyond what is expected between two different datasets of supposedly similar images, leading to very different generalisation scores of the random forest model.

To conclude, the final point is expanded upon. It is reasonable to expect that the datasets that one chooses to train and test on will impart some differences in the scores. However, in the case of these two datasets, it has been shown that the quality of the sets has a very large effect on the precision scores achieved by the machine learning model and researchers should be careful about the datasets used to train and test models. Even though NCT-CRC-HE-100K and CRC-VAL-HE-7K are popular datasets used to train and test machine learning and deep learning models, it seems the quality of the training set is lower than that of the test set, and researchers using these sets to train models should be wary of this fact.

## Data Availability

All data produced are available online at
https://zenodo.org/records/1214456

## References

1. Afshari Mirak, S., et al., N., Mohamed, I.: The growing nationwide radiologist shortage: Current opportunities and ongoing challenges for international medical graduate radiologists. Radiology 314(3), e232625 (Mar 2025)

2. Biau, G., Scornet, E.: A random forest guided tour. TEST 25(2), 197–227 (Jun 2016). 10.1007/s11749-016-0481-7

3. Breiman, L.: Out-of-bag estimation. Tech. rep., University of California Berkeley (1996)

4. Breiman, L.: Random forests. Machine Learning 45(1), 5–32 (Oct 2001). 10.1023/A:1010933404324

5. Breiman, L., et. al: Classification and Regression Trees. Chapman and Hall/CRC, New York (Oct 2017). 10.1201/9781315139470

6. Brito-Pacheco, D., et al.: Relationship between irregularities of the nuclear envelope and mitochondria in hela cells observed with electron microscopy. In: 2024 IEEE ISBI. p. 1–5 (May 2024)

7. Brito-Pacheco, D., Giannopoulos, P., Reyes-Aldasoro, C.C.: Persistent homology in medical image processing: A literature review. medRxiv (Feb 2025). 10.1101/2025.02.21.25322669

8. Edelsbrunner, H., Harer, J.: Persistent homology—a survey, vol. 453, p. 257–282. American Mathematical Society, Providence, Rhode Island (2008). 10.1090/conm/453/08802

9. Ignatov, A., Malivenko, G.: Nct-crc-he: Not all histopathological datasets are equally useful (Sep 2024). 10.48550/arXiv.2409.11546

10. Janitza, S., Hornung, R.: On the overestimation of random forest’s out-of-bag error. PLoS ONE 13(8), e0201904 (Aug 2018). 10.1371/journal.pone.0201904

11. Kather, J.N., et al.: 100,000 histological images of human colorectal cancer and healthy tissue (Apr 2018). 10.5281/zenodo.1214456

12. Kather, J.N., et. al: Predicting survival from colorectal cancer histology slides using deep learning: A retrospective multicenter study. PLOS Medicine 16(1), e1002730 (Jan 2019). 10.1371/journal.pmed.1002730

13. Khvostikov, A., et al.: Tissue type recognition in whole slide histological images. In: Proceedings of the 31th International Conference on Computer Graphics and Vision. Volume 2. p. 496–507. Keldysh Institute of Applied Mathematics (2021). 10.20948/graphicon-2021-3027-496-507

14. Maier-Hein, L., et al.: Why rankings of biomedical image analysis competitions should be interpreted with care. Nat. Commun. 9(1), 5217 (Dec 2018)

15. Project, T.G.: GUDHI User and Reference Manual. GUDHI Editorial Board, 3.11.0 edn. (2025), https://gudhi.inria.fr/doc/3.11.0/

16. Ramos, J., Aung, P.P.: International medical graduates and the shortage of US pathologists: Challenges and opportunities. Arch. Pathol. Lab. Med. 148(6), 735–738 (Jun 2024)

17. Reyes-Aldasoro, C.C.: Multiresolution Volumetric Texture Segmentation. doctoral, University of Warwick (Nov 2004), http://wrap.warwick.ac.uk/67756/

18. Russell, D.K., et al.: Analysis of 2023 cytologists employment survey. J. Am. Soc. Cytopathol. 14(2), 78–85 (Mar 2025)

19. Sun, K., et al.: Automatic classification of histopathology images across multiple cancers based on heterogeneous transfer learning. Diagnostics 13(77), 1277 (Jan 2023). 10.3390/diagnostics13071277

20. van der Walt, S., et. al: scikit-image: image processing in python. PeerJ 2, e453 (Jun 2014). 10.7717/peerj.453

21. Wang, K.S., et al.: Accurate diagnosis of colorectal cancer based on histopathology images using artificial intelligence. BMC medicine 19(1), 76 (Mar 2021). 10.1186/s12916-021-01942-5

